# Right-to-Left Shunt-associated Brain Functional Changes in Migraine: Evidences from a Resting-state FMRI Study

**DOI:** 10.1101/2024.01.23.24301677

**Authors:** Wenfei Cao, Lei Jiao, Huizhong Zhou, Jiaqi Zhong, Nizhuan Wang, Jiajun Yang

**Author notes:** Correspondence: Nizhuan Wang,; Jiajun Yang.

## Abstract

**Background:** Migraine, a neurological disorder under perpetual investigation, has an elusive etiology. An potential association with Right-to-Left Shunt (RLS) exists, yet the precise nature of this connection remains unclear. This study employs the resting-state functional magnetic resonance imaging (rs-fMRI) technique to examine brain functional differences between the migraine patients with and without RLS, aiming at exploring RLS associated alterations in functional segregation and integration.

**Methods:** This study included 32 migraine patients (14 patients with RLS and 18 without RLS), each undergoing rs-fMRI data acquisition. The amplitude of low-frequency fluctuation (ALFF) was employed to investigate functional segregation. Functional connectivity (FC) analysis was conducted to explore the functional integration across distinct brain regions. Graph theory-based network analysis was utilized to assess functional networks in migraine patients with RLS. Pearson correlation analysis further explored the relationship between RLS severity and various functional metrics..

**Results:** Compared with migraine patients without RLS, migraine patients with RLS exhibited a significant increase in the ALFF in the left middle occipital and superior occipital gyrus; As to FC, the reduced connectivity between the left rolandic operculum and the right middle cingulate gyrus was observed in migraine patients with RLS; Based on the brain networks analysis, migraine patients with RLS displayed higher values of the normalized clustering coefficient and greater betweenness centrality in specific regions, including the left precuneus, right insula, and right inferior temporal gyrus. Further, the study found positive correlations between ALFF values in the temporal lobes, thalamus, left middle occipital, and superior occipital gyrus and RLS severity. Conversely, negative correlations emerged between ALFF values in the right inferior frontal gyrus, middle frontal gyrus, and insula and RLS grading. Finally, the study identified a positive correlation between angular gyrus betweenness centrality and RLS severity.

**Conclusion:** RLS-associated brain functional alterations in migraine consisted of local brain regions, connectivity, and networks involved in pain conduction and regulation did exist in migraine with RLS.

## 1. Introduction

In the context of International Classification of Headache Disorders, 3rd edition (ICHD-3) (Arnold M, 2018), migraine is characterized by specific attributes, including unilateral localization, moderate to severe intensity, recurrent episodes, pulsatile headache, and a duration lasting for 4–72 hours. Moreover, it may present associated symptoms such as nausea, vomiting, photophobia, phonophobia, and others. Migraine is a pervasive and debilitating condition that exerts adverse effects on various aspects of individuals’ lives, encompassing marital relationships, parenting responsibilities, emotional well-being, occupational performance, and overall daily health (Buse DC et al., 2019). This intricate disorder can be induced by a variety of factors, including physical exertion, sleep irregularities, dietary choices, meteorological variations, emotional fluctuations, among others (Chądzyński P et al., 2019; Moon HJ et al., 2017; Fernández-de-Las-Peñas C et al., 2017). Nonetheless, the comprehensive pathophysiological underpinnings of migraine remain partially elucidated. An increasing body of research suggests the involvement of various mechanisms, encompassing cortical spreading depression (CSD), vascular alterations, neurogenic inflammation, release of vasoactive substance, disturbances in energy metabolism and genetic factors (Domitrz I et al., 2022; Goadsby PJ et al., 2019; Stanyer EC et al., 2021). In light of this multifaceted backdrop, factors exhibiting correlations with migraine comprise sleep disorders (Holland PR et al., 2018; Kim KM et al., 2019), dysbiosis of gut microbiota (Wen Z et al., 2019; Arzani M et al., 2020), levels of sex hormone (Warnock JK et al., 2017), and genetics (Hautakangas H et al., 2022; Sutherland HG et al., 2019). Furthermore, in recent decades, the association between RLS and migraine has been the subject of debate (Zhao QX et al., 2017; Takagi H et al., 2016).

RLS denotes an anomalous shunting pathway connecting the venous and arterial circulations, with the most prevalent etiology being the patent foramen ovale (PFO), accounting for 95% (Liu K et al., 2020). In clinical assessment of individuals suspected of having RLS, the initial diagnostic step commonly involves the use of contrast-enhanced transcranial doppler (cTCD), recognized for its high sensitivity. A meta-analysis from 2014 reported the sensitivity and specificity of cTCD to be approcimately 97% and 93% respectively (Zhang YS et al., 2021). The association between RLS and migraine stems from the pioneering work of Del et al., who initially disclosed a heightened prevalence of RLS among patients with migraine with aura when compared to healthy individuals (Del Sette M et al., 1998). Subsequent observational studies have consistently supported this association (Lip PZ et al., 2014; Tian DC et al., 2019; Zhao Q et al., 2021; Tang Y et al., 2022). Due to variations in prevalence, a plausible association between RLS and migraine has been suggested. The hypothesis posits that RLS may facilitate the passage of vasoactive substances such as 5-HT, nitric oxide, and kinin directly into the cerebral circulation, potentially triggering migraine attacks (Dalla Volta G et al., 2005; Wilmshurst, P et al., 2006). Moreover, the presence of RLS is implicated in the formation of microemboli, initiating cortical spreading depression (CSD), which, in turn, may activate the trigeminal neurovascular system, leading to headaches (Ashina, M et al., 2019; Sevgi EB et al., 2012).

We speculate that there may be differences in certain brain structures and functions between migration with RLS and without RLS. Recently, more and more neuroimaging evidences demonstrated that distinctions may exist in certain brain structures between individuals experiencing migraines with and without RLS. Specifically, research has focused on white matter hyperintensities (WMH) in cases associating migraine with RLS (Cao W et al., 2022). Several studies have reported notable variations in the prevalence, location, and volume of WMH between migraines with and without RLS (Park HK et al., 2011; Yoon GJ et al., 2012; Iwasaki A et al., 2017). Resting-state functional magnetic resonance imaging (rs-fMRI) is a widely used modality for examining the cerebral functional dynamics of patients under resting conditions. This approach offers notable advantages such as repeatability, ease of operation, and high spatial resolution (Vakamudi K et al., 2012). The analysis of rs-fMRI data encompasses various methods (Zang YF et al., 2012; Wang N et al., 2013; Wang N et al., 2012;Tang XY et al., 2017; Wang Z et al., 2014), broadly categorized into functional segregation and functional integration approaches (Lv H et al., 2018). Functional segregation exemplified by metrics amplitude of low-frequency fluctuation (ALFF), primarily concentrates on assessing the local functional characteristics of specific brain regions (Zou QH et al., 2008; Yu Q et al., 2021). Conversely, functional integration, represented by methods like functional connectivity (FC) and graph theory analysis, delves into the assessment of interregional functional interactions and, essentially, the holistic organization of the brain as an integrated network (Wang N et al., 2017; Wang N et al., 2015; Tang X et al., 2015; Tang XY et al., 2017; Wang N et al., 2018; Yan H et al., 2022; Sporns O 2018; Bullmore E et al., 2009). Numerous rs-fMRI studies have been conducted to elucidate the physiological underpinnings of migraine (Messina R et al., 2022). In particula, aberrant descending pain modulatory pathway prior to the migraine attack, abnormal thalamo-cortical and frontoparietal pathways involved in pain transmission and modulation have been observed in migraine individuals Nie W et al., 2021; Lim M et al., 2021; Mungoven TJ et al., 2022)

In this study, we posit a potential correlation between RLS and functional alterations in integration and segregation in individuals with migraines. To examine this hypothesis, the study meticulously explores RLS-associated brain functional aberrations in migraine individuals with and without RLS The overarching goal is to unravel the underlying connection between RLS and migraines, contributing to a comprehensive understanding of this intricate interplay.

## 2. Material and methods

### 2.1. Participants

The cohort of migraine patients was meticulously assembled from the pool of outpatients seeking medical attention within the Department of Neurology at the Shanghai Sixth People’s Hospital, which is affiliated with the Shanghai Jiao Tong University School of Medicine. All participants provided their explicit informed consent by signing the requisite agreement, signifying their willingness to participate in this study. Moreover, it is important to emphasize that this study received the formal approval of the Ethics Committee at the Shanghai Sixth People’s Hospital Affiliated to Shanghai Jiao Tong University School of Medicine.

In adherence to the inclusion criteria for migraine patients, the following prerequisites were established: (1) Conformance to the diagnostic criteria for migraine as stipulated in ICHD-3; (2) Age range of 14 to 70 years; (3) Inclusion in the study was contingent upon subjects being within the interval between migraine attacks.

Conversely, a series of stringent exclusion criteria were meticulously applied, including: (1) The presence of other primary and secondary headache disorders; (2) Concurrent manifestation of mental illnesses; hypertension, vascular/heart disease, and any major systemic disorders; (3) A history of substance addiction involving alcohol or drugs; (4) The presence of contraindications that rendered individuals unsuitable for MRI or cTCD examinations.

All the participants provided written informed consent to participate in the current study. The study was registered with Clinical Trial (ChiCTR2300067636) and obtained ethical approval from the Ethics Committee of Shanghai Sixth People’s Hospital Affiliated to Shanghai Jiao Tong University School of Medicine (Approval No. 2022-KY-194(K)).

### 2.2 Data Acquisition

#### 2.2.1. Demography and Clinical Data

Demographic data, comprising age, gender, body mass index (BMI), and educational history were meticulously acquired. Furthermore, a structured diagnostic interview was conducted to elicit comprehensive information concerning the patients’ symptoms, including the presence or absence of migraine triggers, aura phenomena, the frequency and duration of migraine episodes, remission and exacerbation factors, and any concomitant symptoms experienced. Notably, a visual analog scale (VAS) was employed to quantitatively gauge the severity of pain reported by the patients.

#### 2.2.2 Contrast-enhanced Transcranial Doppler

All participants underwent a cTCD examination, administered by skilled physicians to ascertain the presence of RLS. During this examination, the patients reclined in a supine position, and a three-way catheter was inserted into the left elbow vein to establish a venous access. Two 20 mL syringes were connected to the catheter, one containing a solution comprising 8 mL of physiological saline, 1 mL of air, and 1 mL of autologous blood. The contents of the two syringes were then rapidly interchanged to generate an air microbubble suspension. Subsequently, this microbubble suspension was promptly injected into the elbow vein. Simultaneously, ultrasonic monitoring of microembolic signals in the middle cerebral artery (MCA) was conducted using a transcranial Doppler ultrasound probe via the temporal bone window, with recordings lasting for 20 seconds. This examination was performed both at rest and during the Valsalva maneuver. The RLS was graded based on the maximum number of microemboli observed. Specifically, if 1-10 microbubbles (one side) were detected, it was categorized as a small RLS; if 11-20 microbubbles (one side) were observed, it was classified as a medium RLS. Moreover, if more than 20 microbubbles (one side) or curtain patterns were noted, it was designated as a large RLS (Zhang YS et al., 2021).

#### 2.2.3 FMRI Data Acquisition

Initially, each subject underwent a comprehensive brain MRI examination encompassing various sequences, such as T1-weighted images (T1WI), T2-weighted images (T2WI), fluid attenuated inversion recovery (FLAIR) images, and diffusion-weighted imaging (DWI) images, with the final diagnosis being conferred by a qualified medical specialist. Any data presenting secondary intracranial lesions were subsequently excluded from the analysis, while fMRI data acquisition proceeded for those without such lesions. All MRI and fMRI scans were executed employing a 3.0T GE MRI scanner (SIGNA, GE Healthcare). Furthermore, participants were instructed to remain in an awakened state with their head securely immobilized, maintaining physical relaxation, and refraining from intentional cognitive activity throughout an 8-minute resting-state fMRI session.

The resting-state fMRI parameters of all participants in this study adhered to the following standards: repetition time (TR) =3000 ms, echo time (TE) =30 ms, field of view (FOV) =240 mm×240 mm, matrix =128×128, slice thickness =4 mm, flip angle =90°, comprising 38 axial slices arranged in parallel, with 160 time points acquired.

### 2.3. Data Preprocessing

Preceding data analysis, a sequence of preprocessing procedures and stringent quality control measures were meticulously executed on rs-fMRI data. These tasks were diligently carried out using the Data Processing and Analysis of Brain Imaging (DPABI) toolbox (version 6.1, http://rfmri.org/dpabi) based on MATLAB (The Math

Works, Natick, MA, USA) software, to remove the effects of data acquisition, physiological noise, and individualized variations in the subject’s brain, as well as to ensure the confidence and sensitivity of the group-level analysis (Yan C et al.; 2016). Figure. 1 specifies the detailed steps of preprocessing and its role.

**Fig. 1.**
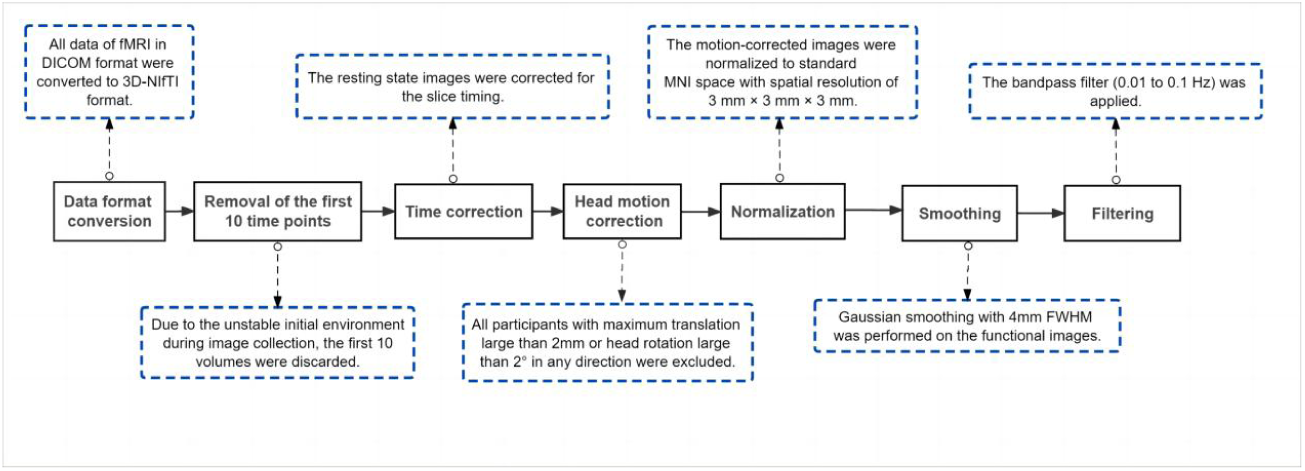
Procedures of preprocessing. DICOM: Digital Imaging and Communications in Medicine. NIfTI: Neuroimaging Informatics Technology Initiative. MNI: Montreal Neurological Institute. FWHM: Full Width Half Maximum.

### 2.4. ALFF Analysis

Preceding the application of filtering, the ALFF was computed using the DPABI software (Yan C et al., 2016; Yan C et al., 2010), relying on the preprocessed dataset. This entailed the transformation of the time series data of an individual voxel into a frequency spectrum by means of the Fourier transform. Subsequently, the summation of amplitudes within the frequency range of 0.01-0.1 Hz was executed to derive the ALFF (Lv H et al., 2018; Zuo XN et al., 2010; Biswal B et al., 1995). This procedure was systematically applied to all voxels throughout the entire brain, thereby producing the comprehensive ALFF map representing the entire cerebral structure of the subject.

### 2.5. Network Construction and Analysis

Brain network construction and subsequent analysis were executed using the DPABI NET (version 1.1) toolbox. The anatomical automatic labeling (AAL) template (available at: http://www.gin.cnrs.fr/en/tools/aal/), a widely recognized brain atlas, was adopted to partition the entire brain into a total of 116 distinct brain regions (Tzourio-Mazoyer N et al., 2002). It is worth noting that 26 of these regions are located within the cerebellum. Consequently, only the 90 regions of interest (ROIs) within the cerebral cortex were chosen as nodes for network construction. The average time series of each ROI were extracted, and the Pearson’s correlation coefficient between the average time series of each two ROIs was calculated, thus obtaining a 90×90 two-dimensional matrix to establish the brain functional network. Additionally, a range of sparsity settings, varying from 0.01 to 0.5, were applied to construct a brain-weighted network. This range ensured that a connection matrix involving all 90 nodes was encompassed within this specified sparsity range.

Complex-network (graph) analysis was performed using Brain Connectivity Toolbox (https://www.nitrc.org/projects/bct/) (Rubinov M et al., 2010) while the BrainNet viewer (https://www.nitrc.org/projects/bnv/) was utilized to display results (Xia M et al., 2013). Brain network topology analysis includes “small world” properties as well as node properties. In line with the prior literature (Yan CG et al., 2013), we computed each small-world property’s value across 50 different sparsity levels. Furthermore, we determined the area under the curve (AUC) for each small-world metric within the sparsity range of 0.01-0.34, allowing the more sensitive alteration detection in small-world network of brain. Subsequently, we derived the values for the following small-world properties at each sparsity level: characteristic shortest path length (L_p_), clustering coefficient (C_p_), normalized clustering coefficient (γ), normalized characteristic shortest path length (λ), small-worldness (σ), local efficiency (E_loc_), global efficiency (E_glob_), degree centrality, betweenness centrality, and nodal efficiency (Yang H et al., 2021).

### 2.6. Statistical Analysis

Demographic characteristics, including age, gender, and years of education, and clinical features such as migraine duration, frequency, and VAS score, underwent rigorous statistical analysis utilizing Statistical Product and Service Solutions (SPSS) software (https://www.ibm.com/spss). Categorical variables were assessed with chi-square tests, and numerical variables were statistically evaluated through independent samples t-tests and non-parametric tests. A significance level of p < 0.05 indicated statistical significance.

Two-sample t-tests were conducted for ALFF in both groups, utilizing age and gender as covariates. In order to minimize the false-positive rates, a stringent correction for multiple comparison was applied at the clump level (voxel p < 0.001, cluster p-value < 0.05, corrected using GFR) in this study. The present study reveals the surviving corrected clumps. ALFF values were extracted from the migraine with RLS group for subsequent Pearson’s linear correlation analysis with RLS grading. The statistical threshold was established at p < 0.05, with a requirement of a cluster size exceeding 60.

Two-sample t-tests were carried out to assess functional connectivity differences between the two groups, with a significance threshold set at p < 0.001. Furthermore, small-world properties (with a significance threshold at p<0.05) and nodal properties (with a significance threshold at p<0.01) were subjected to two sample t-tests, while accounting for age and gender as covariates. The surviving nodes were then reported. In all Pearson’s linear correlation analyses in the current study involving RLS grading, the significance level was set at p < 0.05.

## 3. Results

### 3.1 General Characteristics and Clinical Features

The clinical and demographic data of all patients are presented in Table 1. No significant differences were observed between the two groups concerning gender, age, years of education, duration of migraine (years), frequency (in times per month) and

**Table 1.** Characteristics and clinical profiles of migraine patients with and without RLS.

| Characteristics | Migraine |  | P value |
| --- | --- | --- | --- |
|  | With RLS (n=14) | Without RLS (n=18) |  |
| Sex, n% |  |  |  |
| Male | 3 (21.43%) | 1 (5.56%) |  |
| Female | 11 (78.57) | 17 (94.44%) |  |
| Age (years) | 36.71 ± 6.62 | 41.56 ± 12.41 | 0.168 |
| Years of education | 13.71 ± 2.37 | 13.11 ± 4.01 | 0.889 |
| Migraine duration (years) | 12.71 ± 9.45 | 16.5 ± 12.49 | 0.401 |
| Frequency (times/month) | 3.69 ± 3.24 | 1.92 ± 1.46 | 0.086 |
| VAS | 8.11 ± 0.92 | 7.83 ± 1.83 | 0.969 |

RLS: Right-to-Left Shunt; VAS: Visual Analogue Scale

VAS scores. In total, 32 patients were enrolled in this study, comprising 14 migraine patients with RLS and 18 migraine patients without RLS. Within the RLS group, five patients with large RLS, three had moderate RLS, and six showed mild RLS.

### 3.2. Differences in ALFF

The results demonstrated notably elevated ALFF in Occipital_Sup_L/Occipital_Mid_L (AAL3) regions ALFF in migraine patients with RLS in comparison to migraine patients without RLS (voxel p < 0.001, cluster p-value < 0.05, GFR corrected). Please refer to Table 2 and Fig. 2.

**Fig. 2.**
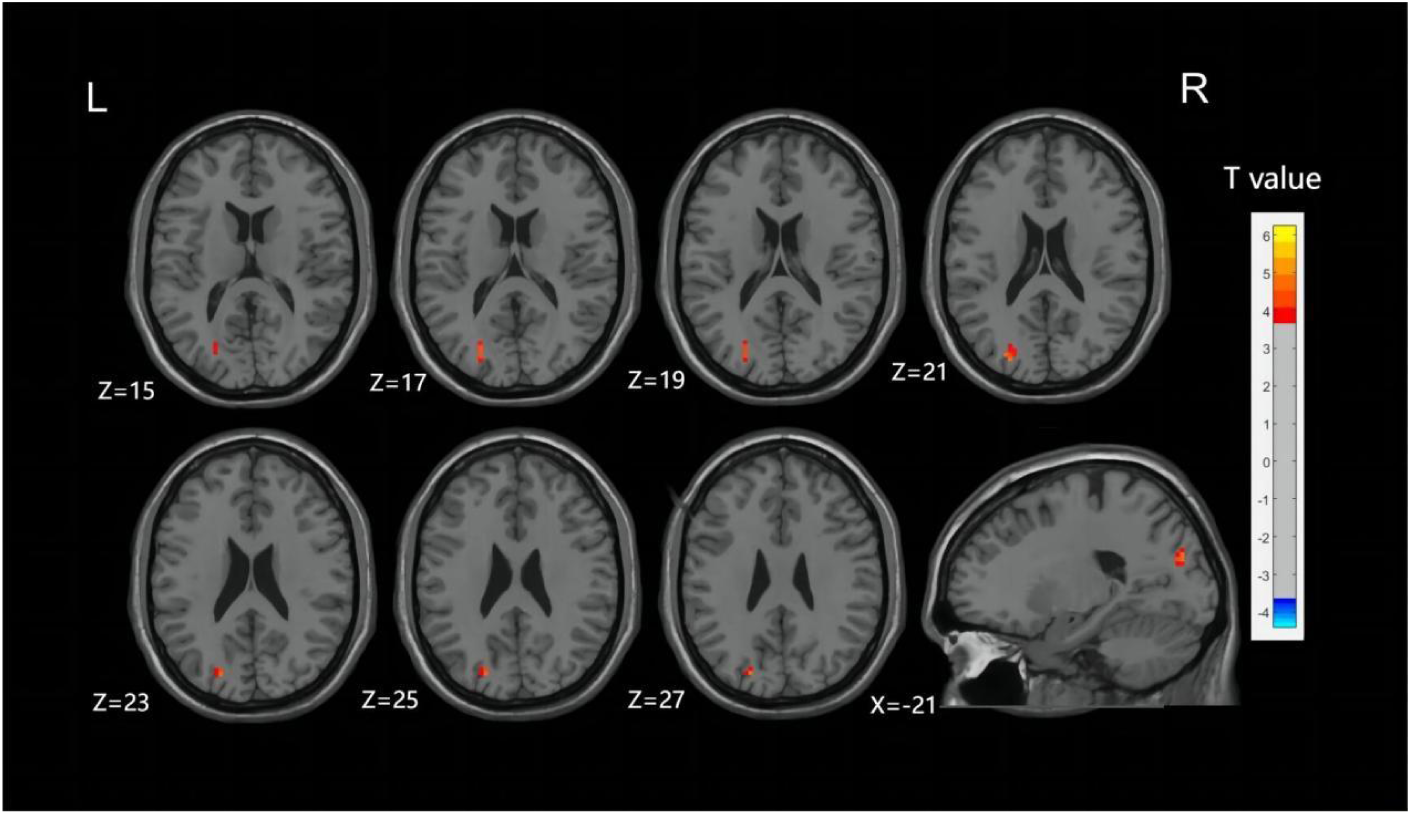
Differences in ALFF between migraine patients with and without RLS. The color red denotes increased ALFF in migraine patients with RLS compared to those without RLS. The corresponding color bar denotes the t-value. R: right; L: left.

**Table 2.** Brain regions with significant ALFF differences between migraine patients with and without RLS.

| Brain regions (AAL3) | Cluster size | Cluster volume | MNI coordinates | Peak T |
| --- | --- | --- | --- | --- |

|  | (voxels) | (mm<sup>3</sup>) | (mm) |  |  | value |
| --- | --- | --- | --- | --- | --- | --- |
|  |  |  | X | Y | Z |  |
| Occipital_Sup_L | 23 | 621 | -21 | -81 | 27 | 4.83 |
| Occipital_Mid_L |  |  |  |  |  |  |

MNI: Montreal Neurological Institute; AAL: Anatomical Automatic Labeling.

### 3.3. Correlation of ALFF with RLS Degree

As illustrated in Fig. 3 and detailed Table 3, specific functional brain regions exhibited notable correlations with RLS classification. The bilateral temporal lobes (AAL3: Hippocampus_R, ParaHippocampal_R, Hippocampus_L, Temporal_Inf_L, Temporal_Sup_L), bilateral thalamus (AAL3: Thal_VL_R, Thal_VPL_R, Thal_VA_R, Thal_VL_L, Thal_MDm_L, Thal_IL_L) and left occipital lobe (AAL3: Occipital_Mid_L, Occipital_Inf_L) displayed a significant positive correlation between ALFF and RLS classification (p < 0.05, number of cluster > 60). In contrast, ALFF in regions of right inferior frontal gyrus (AAL3: Frontal_Inf_Oper_R, Frontal_Inf_Tri_R, Frontal_Inf_Orb_2_R), bilateral middle frontal gyrus (AAL3: frontal_Mid_2_R, frontal_Mid_2_L) and bilateral insula (AAL3: Insula_L, Insula_R) exhibited a significant negative correlation with RLS classification (p < 0.05, number of cluster > 60).

**Fig. 3.**
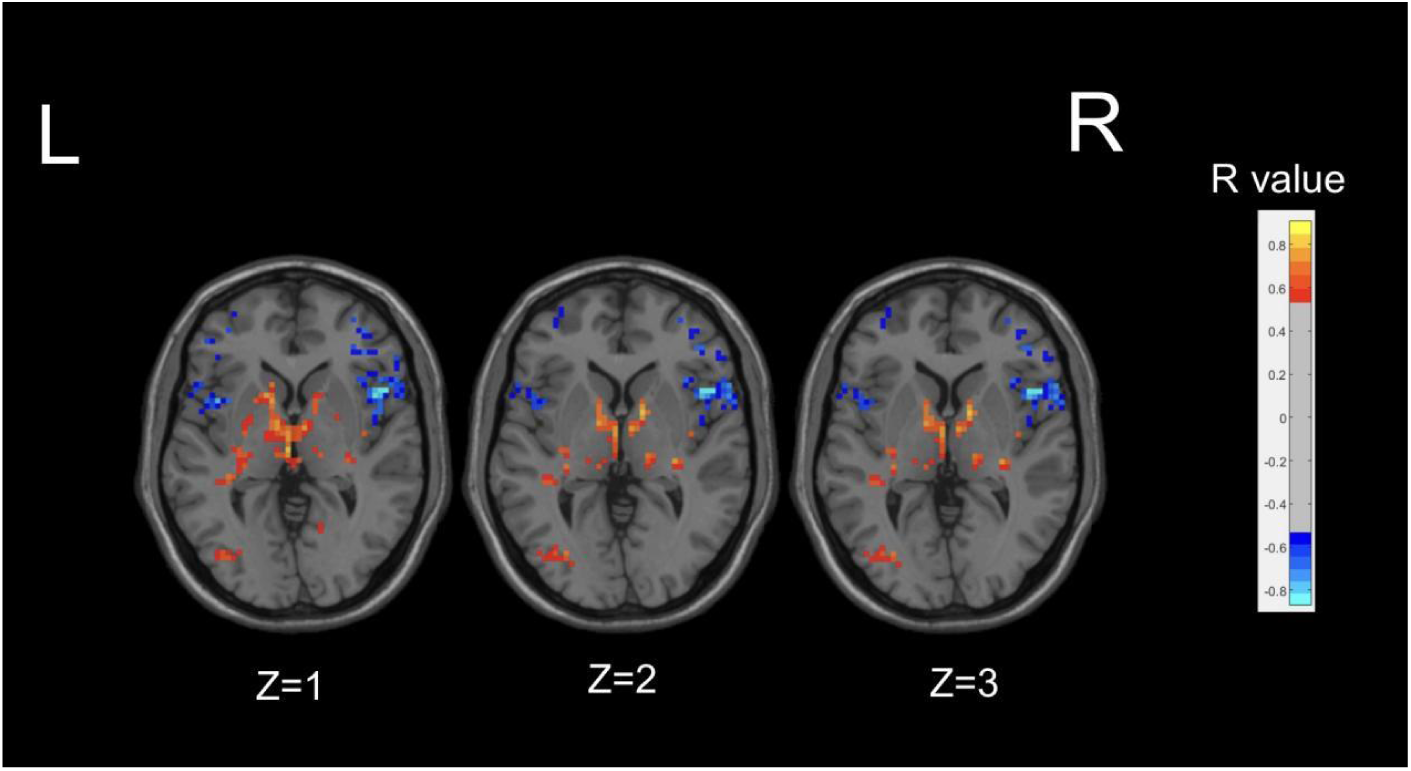
Correlation maps illustrating the relationship between ALFF and RLS degree in migraine patients with RLS. Warm colors, such as red, denote a positive correlation, while cool colors, like blue, signify a negative correlation. The color bar denotes the R-value. R: right; L: lef.t

**Table 3.**
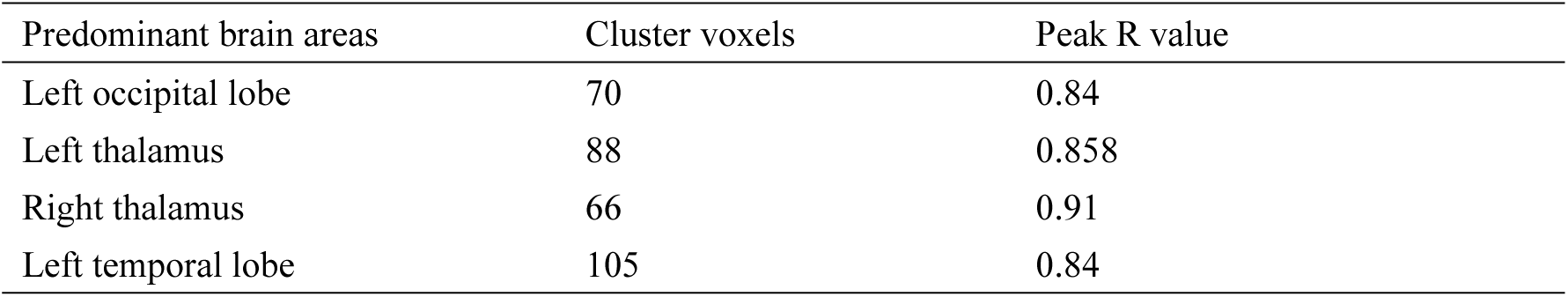
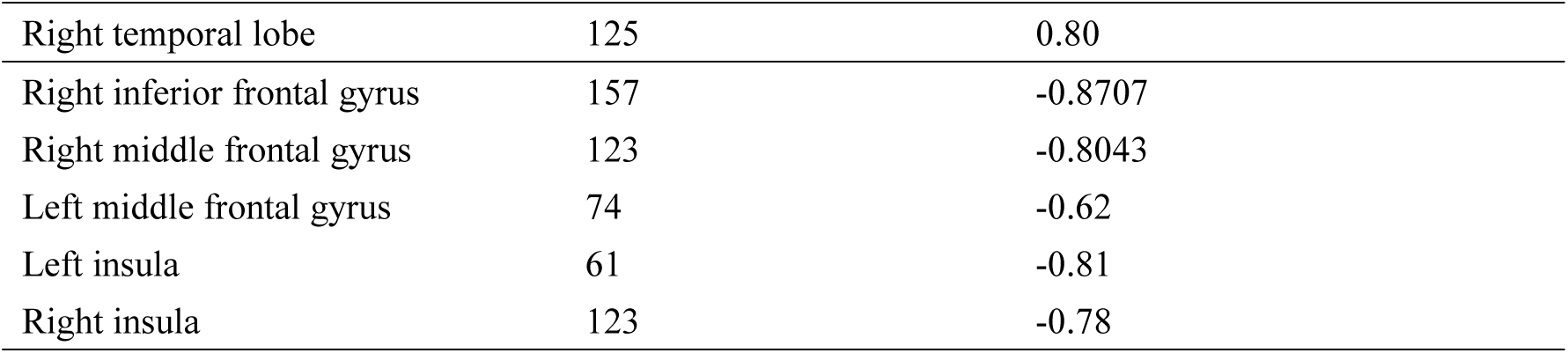
Correlation coefficients representing the relationship between ALFF and RLS degree in migraine patients with RLS.

| Predominant brain areas | Cluster voxels | Peak R value |
| --- | --- | --- |
| Left occipital lobe | 70 | 0.84 |
| Left thalamus | 88 | 0.858 |
| Right thalamus | 66 | 0.91 |
| Left temporal lobe | 105 | 0.84 |
| Right temporal lobe | 125 | 0.80 |
| Right inferior frontal gyrus | 157 | -0.8707 |
| Right middle frontal gyrus | 123 | -0.8043 |
| Left middle frontal gyrus | 74 | -0.62 |
| Left insula | 61 | -0.81 |
| Right insula | 123 | -0.78 |

The preceding findings highlight the significance of ALFF differences between the two groups, with the most prominent distinctions observed in Occipital_Sup_L and Occipital_Mid_L (AAL3). Furthermore, the ALFF values in Occipital_Mid_L and Occipital_Inf_L (ALL3) exhibited a positive correlation with RLS classification. This may imply a substantial impact of RLS on the occipital lobe. To further corroborate this observation, we extracted the ALFF values from these distinct brain regions (Occipital_Sup_L and Occipital_Mid_L) in migraine patients with RLS and assessed their correlation with RLS grading. The results, illustrated in Fig. 4, indicated a positive correlation between the ALFF values of the Occipital_Sup_L and Occipital_Mid_L regions and RLS grading, with an R-value of 0.542 (p < 0.05).

**Fig. 4.**
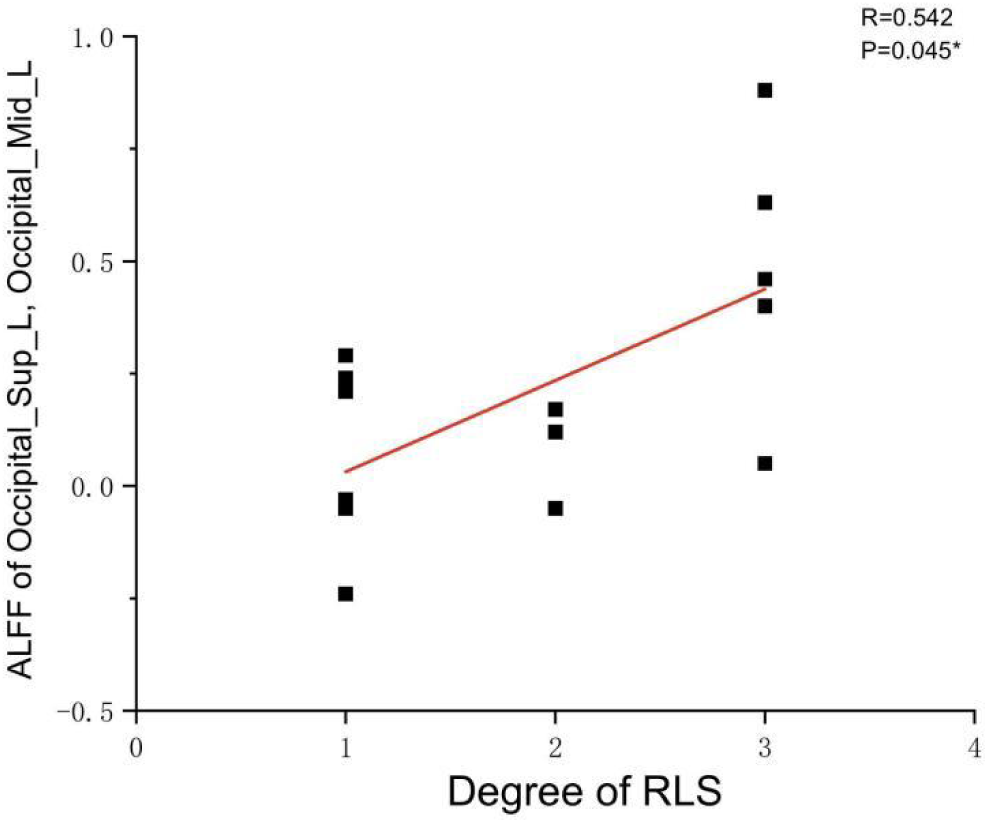
Correlation illustration between ALFF values in Occipital_Sup_L, Occipital_Mid_L regions and RLS degree. Each black dot denotes a sample.*P<0.05. ALFF: Amplitude of Low-frequency Fluctuation; RLS: Right to Left Shunt.

### 3.4. Functional Connectivity

Illustrated in Fig. 5, the migraine group with RLS exhibited a noteworthy reduction in functional connectivity within Roland_Opper_L and Cingulum_Mid_R (AAL), when compared to the migraine group without RLS (p < 0.001).

**Fig. 5.**
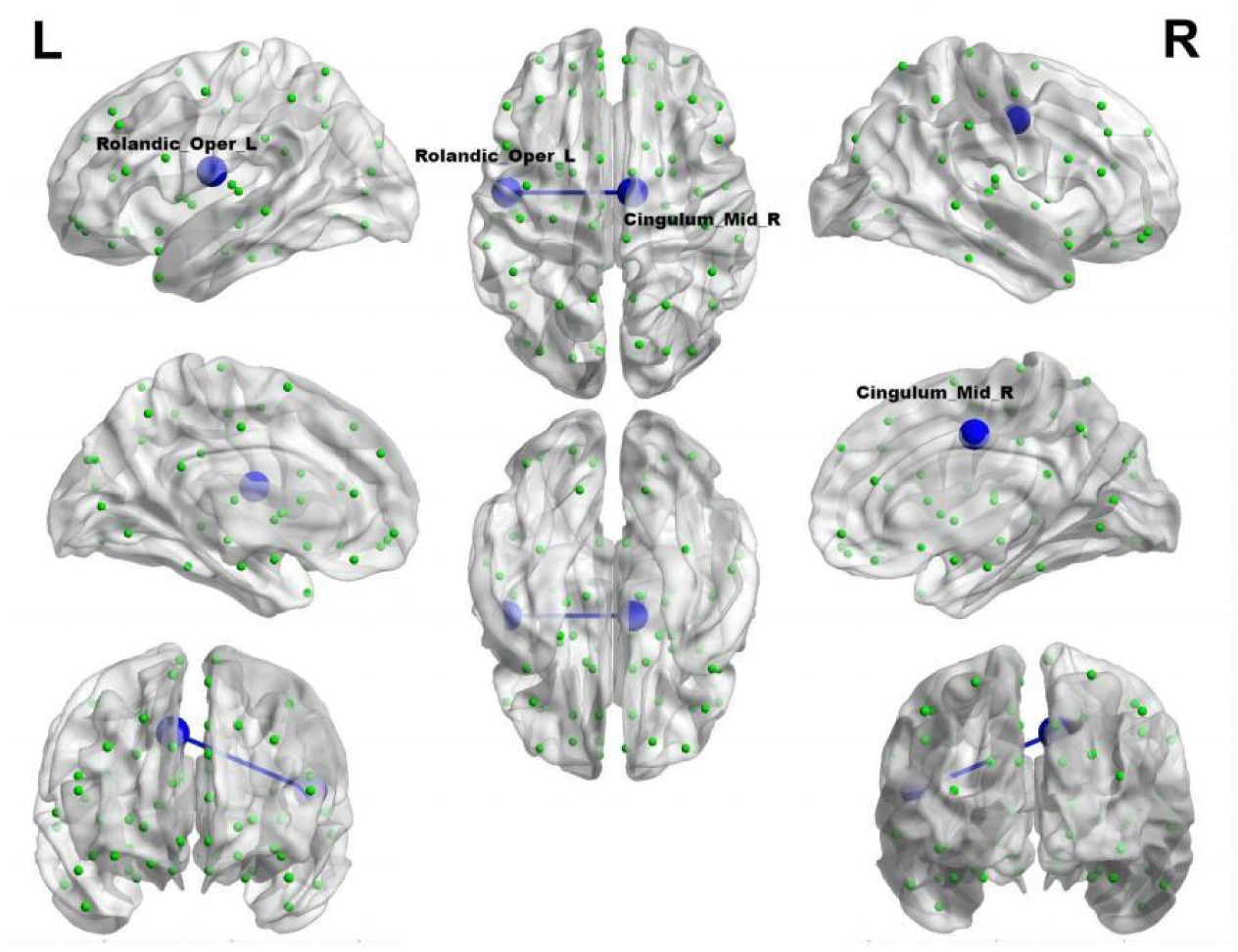
Reduced connectivity between Roland_Opper_L and Cingulum_Mid_R of the migraine group with RLS in comparison to the migraine group without RLS. The azure sphere symbolizes a node corresponding to a specific brain region, while the blue line signifies a decline in functional connectivity. R: right; L: left.

### 3.5. Small-world Property

The analyses of small-world properties revealed that both sets of data displayed σ greater than 1 and exhibited C_p_ values (λ close to 1) similar to the randomized network, as shown in Fig. 6. Consequently, both groups exhibited small-world properties in their functional brain networks. Furthermore, the AUC of γ in migraine patients with RLS group was significantly higher than that of the group without RLS (p < 0.05), as demonstrated in Fig. 7. However, the remaining indices, including σ AUC, λ AUC, E_glob_, E_loc_, C_p_ and L_p_, did not exhibit significant difference between the two groups.

**Fig. 6.**
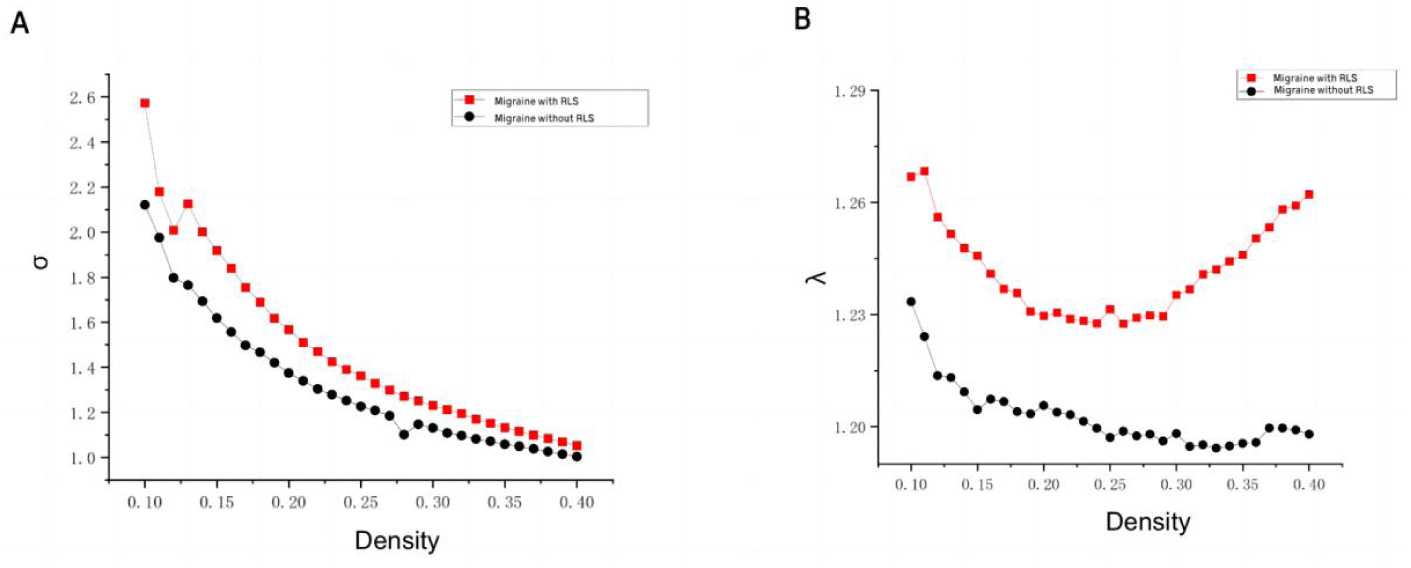
Comparison of “Small World” properties between two groups. **A:** A comparison of “σ” property between two groups. **B:** A comparison of “λ” property between two groups. The typical small-world network architectures, characterized by λ ≈ 1 and σ > 1, were observed across different sparsity levels. Black lines represent the migraine group without RLS, while red lines represent the migraine group with RLS. No statistically significant differences were identified between the two groups.

**Fig. 7.**
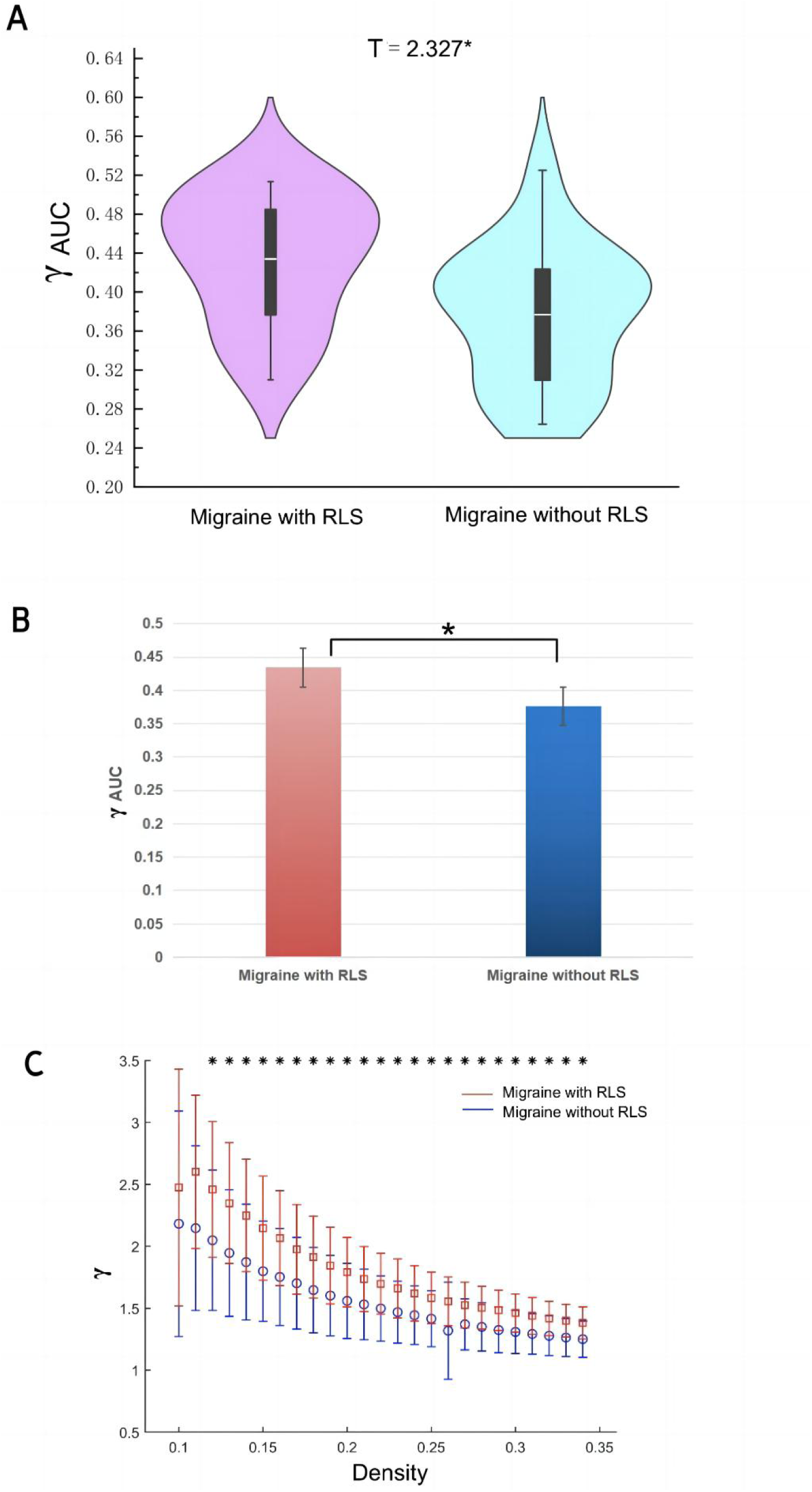
Differences in network topological properties between migraine patients with and without RLS. **A** Violin plots depict the distribution of mean γ AUC values, highlighting the contrast between migraine with RLS and without RLS. **B** The bar graph displays the significant AUC values of γ between the two groups. **C** γ values are shown across a density range spanning from 10% to 34%. Each point, accompanied by an error bar, represents the mean and standard deviation at specific density levels, respectively. * denotes significant differences.

### 3.6. Nodal Properties

In comparison to control subjects, the migraine group with RLS exhibited significantly elevated nodal Betweenness centrality in the Insula_R, Precuneus_L, and Temporal_Inf_R (ALL) (p < 0.01), as demonstrated in Fig. 8. However, no significant disparities were found in nodal degree and nodal efficiency between the two groups.

**Fig. 8.**
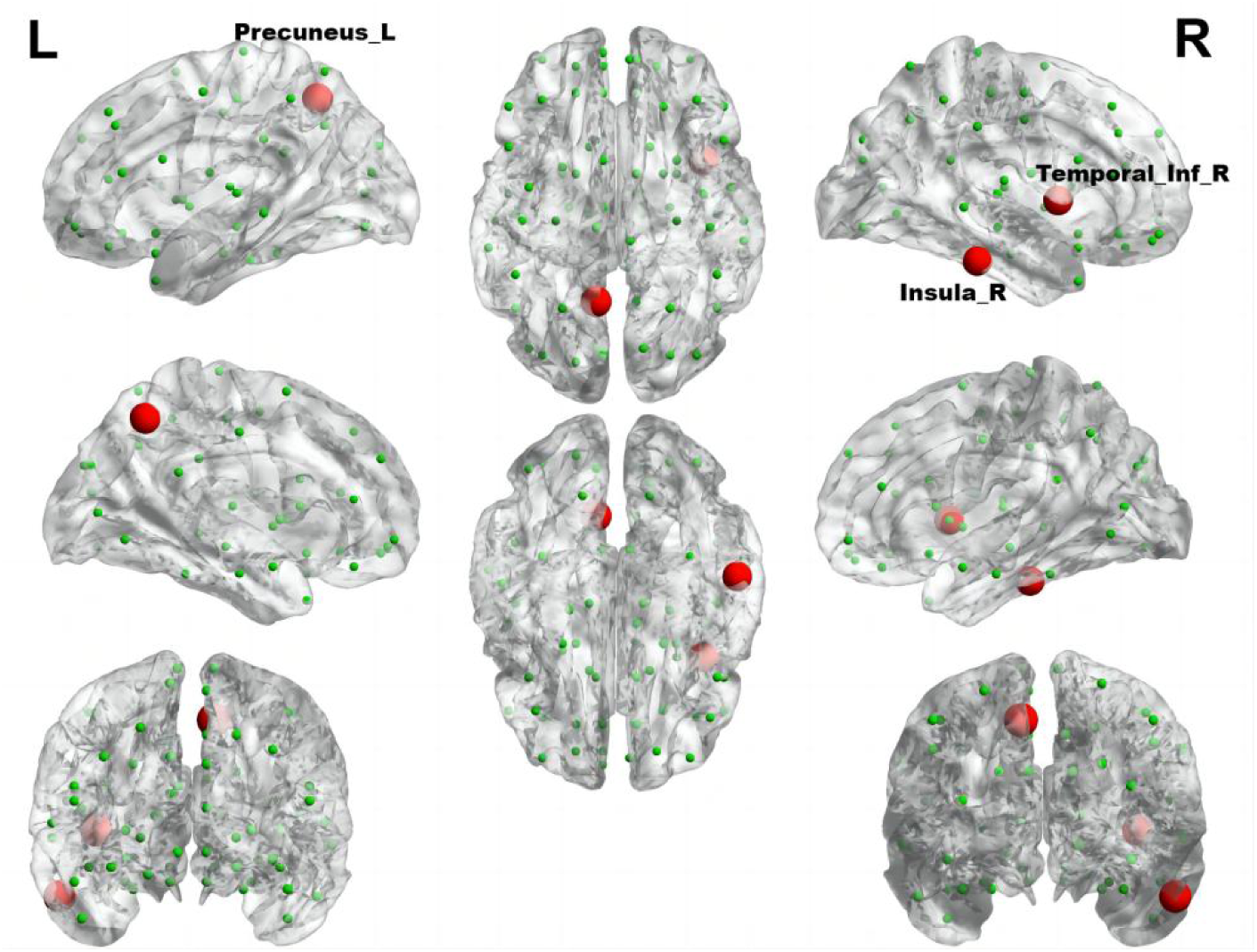
The brain regions with statistically significant difference in Betweenness centrality between the migraine groups with RLS and without RLS. The red spheres represent the brain nodes with increased betweenness centrality in migraine with RLS compared to migraine without RLS. R: right; L: left.

Using RLS grading as the variable, and accounting for age and head movement coefficients as covariates, we employed the DPABI NET toolbox to investigate potential correlations between the brain topological metrics and RLS grading within the migraine patients with RLS group (p < 0.05). The results demonstrated that RLS grading is positively correlated with the betweenness centrality of Angular_R (AAL) (p < 0.01) (Fig. 9). However, none of the seven topologic small-world parameters including σ AUC, Ƴ AUC, λ AUC, L_p_ AUC, C_p_ AUC, E_glob_ AUC, and E_loc_ AUC exhibited a significant correlation with RLS grading. Furthermore, no noteworthy correlation was observed between nodal degree and RLS grading.

**Fig. 9.**
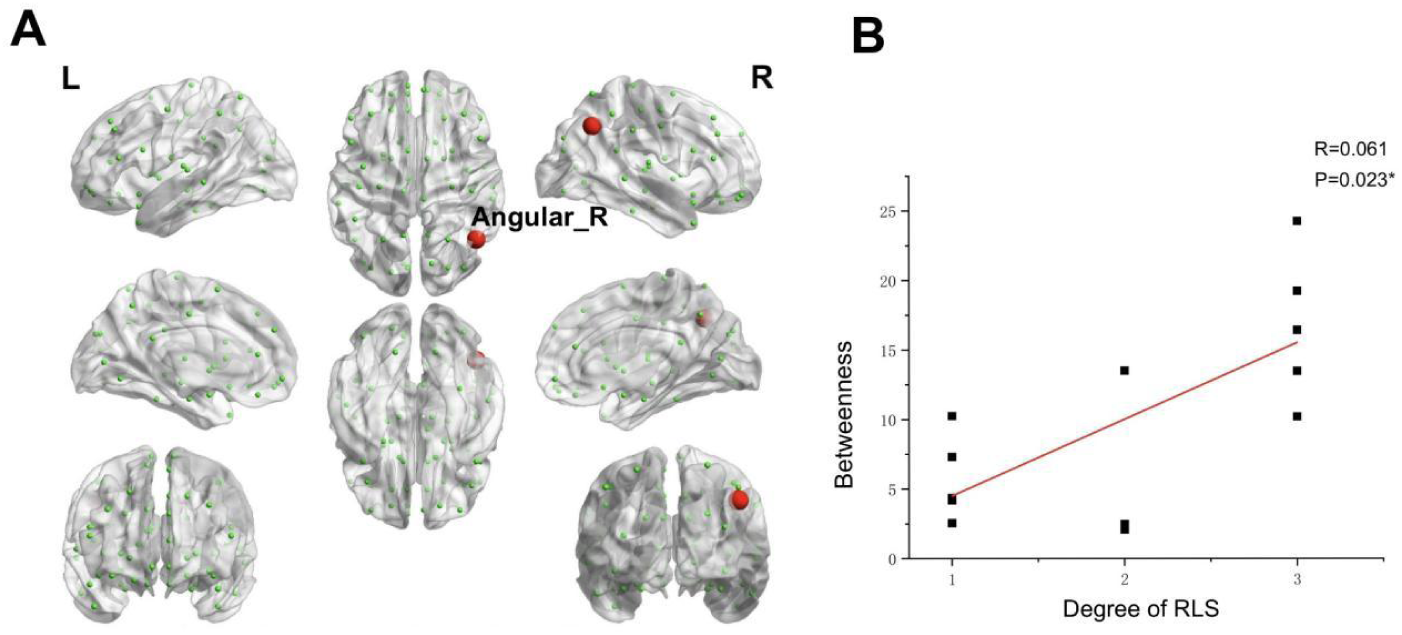
Correlations between the betweenness centrality and RLS grading. **A:** Brain map depicting node with positive correlation. Red node represents those demonstrating a correlation between betweenness centrality and RLS grading. Each black dot signifies an individual sample. **B:** Correlation graph. *P < 0.05. RLS: Right-to Left Shunt.

## 4. Discussion

This pioneering study systematically investigates functional differences in the brain, focusing on regional function, interregional connectivity, and complex network properties in individuals with migraines, both with and without RLS, utilizing rs-fMRI. Our primary objective is to assess RLS-related functional alterations at both localized and global levels in individuals with migraines and RLS. Our findings indicate a significant increase in ALFF, particularly in the left middle occipital gyrus and superior occipital gyrus, within the RLS-positive group, with a positive correlation observed with RLS severity. Additionally, we note a noteworthy reduction in functional connectivity between the left rolandic operculum and the right middle cingulate gyrus in the RLS-positive group.

Employing graph-theoretic analysis, we determined that while both groups of migraine patients exhibited small-world network properties across the entire brain, the RLS-positive group displayed significantly higher values of γ. This suggests that certain nodal properties may have been locally altered. Correspondingly, significant differences in nodal betweenness centrality were identified between the two groups. Notably, right insula, left precuneus, and right temporal regions exhibited notably higher nodal betweenness centrality in migraine patients with RLS compared to those without RLS. Additionally, the betweenness centrality of the right angular gyrus exhibited a positive correlation with RLS severity.

### 4.1. ALFF Difference between Two Groups

ALFF, reflecting the intrinsic neural activity of the brain, serves as a valuable tool for assessing the functional attributes of specific disease-associated brain regions (Zang YF et al., 2007; Liu L et al., 2020; Wang S et al., 2023; Zhe X et al., 2023;). Elevated ALFF values signify heightened spontaneous neural activity in the brain area, while reduced ALFF values indicate decreased spontaneous activity. A recent investigation reported an increased detection rate of RLS in individuals experiencing migraines with visual aura, underscoring a robust correlation between RLS and visual aura (Khessali H et al., 2015). Furthermore, Tu Y et al. previously identified substantial functional connectivity anomalies involving the occipital cortex, precuneus, and thalamus in migraine patients through dynamic analysis of brain functional connectivity (Tu Y et al., 2019). In alignment with our current study, we uncovered augmented spontaneous activity in the middle occipital and superior occipital gyrus among migraine patients with RLS in comparison to those without RLS, with a notable correlation observed concerning the severity of RLS. Hence, we posit that the accumulation of microemboli and vasoactive substances, engendered by RLS, within the occipital lobe may be contributing to the observed alterations in spontaneous brain activity. This phenomenon may also elucidate the heightened likelihood of visual aura manifestation in patients with RLS.

The pain transmission pathway comprises three neuronal levels: peripheral primary neurons initially detect injurious stimuli and relay them via afferent fibers to interneurons. Subsequently, this information is conveyed through the spinothalamic tract and spinoreticular tract to tertiary neurons residing in the thalamus, the central sensory hub. Ultimately, the thalamus transmits the noxious signals to various regions of the cerebral cortex, including the bilateral insulae, prefrontal cortex, anterior cingulate cortex, and somatosensory cortex (Liu J et al., 2012; Baron R et al., 2010). The trigeminal vascular theory, stemming from this foundational model, currently serves as the predominant framework for investigating migraine mechanisms (Lee GI et al.,2020). At its core, this theory hinges on a three-tiered neuronal mechanism, encompassing the trigeminal ganglia (primary neurons), brainstem neurons (intermediate neurons), and thalamus (tertiary neurons). Ultimately, the thalamus dispatches nociceptive information to various regions within the cerebral cortex, including the auditory cortex, olfactory cortex, insular cortex, primary motor cortex, secondary motor cortex, periaqueductal gray matter, posterior subcortical cortex, parietal cortex, primary somatosensory cortex, secondary somatosensory cortex, primary visual cortex, secondary visual cortex, and more. This intricate interplay generates the sensation of pain (Khan J et al., 2021). In our present investigation, we observed significant correlations between RLS grading and ALFF in the thalamus, as well as the occipital, prefrontal, and insular cortex regions. Importantly, several of these associations exhibited positive correlations. These findings signify that RLS may influences the pain transmission pathways, thereby reinforcing the association between RLS and migraine.

### 4.2. Functional Connectivity Difference between Two groups

The rostral anterior cingulate cortex assumes a pivotal role in pain modulation, serving as a linchpin in the downstream pain inhibitory system. Li and colleagues have previously reported a reduced resting-state functional connectivity between the rodent anterior cingulate cortex (rACC)/medial prefrontal cortex (mPFC) and periaqueductal grey (PAG) in migraine patients compared to healthy controls (Ashina M et al., 2019). In alignment with these findings, our results also indicate decreased connectivity involving the central sulcus and the medial and paracingulate cingulate gyrus in RLS-positive group compared with RLS-negative group. In addition, the cingulate gyrus primarily governs the regulation of visceral functions, potentially elucidating the common presence of nausea and vomiting symptoms alongside migraine, attributed to the abnormal visceral regulation orchestrated by the cingulate gyrus (Li Z et al.,2016). Thus, we speculate that RLS may have a potential impact on the pain regulatory pathway in migraine patients.

### 4.3. Difference in Small-World Properties and Nodal Properties

Graph theory analysis is increasingly applied in migraine research, with several studies elucidating the small-world properties of functional brain networks in various migraine conditions, including migraine without aura and chronic migraine (Liu J et al., 2012; Li K et al., 2017; Dai L et al., 2021). Our investigation, too, revealed that both migraine groups, with and without RLS, exhibited small-world properties within their functional brain networks. However, substantial distinctions in topological properties remained evident between the two cohorts. Notably, the RLS-positive group displayed higher γ values, indicative of intensified small-world properties and enhanced efficiency in brain network message transmission. This heightened efficiency sustains an optimal equilibrium between functional segregation and integration. It is plausible that these differences in network topology are attributable to RLS, fostering a more streamlined information exchange within the brain network. Consequently, this refined network may perceptively and expeditiously transmit pain signals among pain-related brain regions.

Betweenness centrality, an index signifying local network attributes, gauges a node’s influence on information transmission amid other nodes. While we have scrutinized the overall small-world characteristics of the brain network in migraine patients with RLS, it remains imperative to delve deeper into the potential alterations within specific local nodes responsible for information transfer in the functional brain network, possibly influenced by RLS. In contrast to the RLS-negative group, our study detected heightened betweenness centrality in the group afflicted by RLS. The insula’s pivotal role in triggering the pain matrix network is well-documented (Labrakakis C 2023). Functioning as a hub within the cortex, the insula orchestrates complex sensations and emotions associated with migraine symptoms. This is accomplished through intricate interconnections linking the frontal, temporal, and parietal cortices, basal ganglia, thalamus, and limbic structures (Borsook D, et al.,2016). Moreover, the inferior temporal gyrus plays a fundamental role in the assembly of the ventral visual pathway, primarily responsible for shaping visual content (Li Q et al., 2018). Our study revealed that RLS-positive group exhibited significantly elevated betweenness centrality in this region compared to RLS-negative group. Thus, it is plausible that RLS’s influence extends to the inferior temporal gyrus, rendering migraine patients with RLS more susceptible to visual aura. This aligns with prior research by Kijima Y et al., wherein they reported higher RLS grades in the migraine group with frequent visual aura compared to those with episodic visual aura or no visual aura (Kijima Y et al., 2015). Consequently, our study further supports the association between RLS and migraine, viewed through the lens of brain function.

The angular gyrus, positioned within the parietal lobe, emerges as a pivotal structure implicated in the intricate process of multisensory integration. It serves as a central hub orchestrating the harmonious synthesis of information from diverse sensory modalities across the cerebral landscape (Dong L et al., 2023; de Pasquale F et al., 2012). Our comprehensive data analysis, notably, illuminated a conspicuous and statistically significant correlation between the betweenness centrality of the angular gyrus and the grading of RLS. This observation implies a possible association between RLS and sensory integration in migraine.

### 4.4 Limitations and Future Work

This study warrants acknowledgment of certain potential limitations. To commence, the relatively modest sample sizes within both study groups may introduce constraints on the precision and generalizability of the findings. Secondly, the lack of a control group without migraine but with and without a RLS prevents us from detecting subtler changes related to RLS in migraine patients. Future research endeavors should consider broader subject recruitment, encompassing a more extensive cohort of participants, including healthy controls. We should further incorporate healthy control with and without RLS, to explore whether there is an effect of RLS on brain function, which may induce migraine attacks. This will help us to verify whether RLS is a risk factor for migraine. Thirdly, because of the absence of the information of heartbeat and respiration during rs-fMRI scans, the potential confounding effect of heartbeat and respiration were not regressed. Heartbeat and respiratory signals should be collected in future research to make the results more rigorous. Fourthly, it is essential to recognize that this study adhered to a cross-sectional design. Consequently, it cannot elucidate the causal dynamics underpinning the observed aberrations in brain functional imaging. In other words, it remains uncertain whether these anomalies precede the onset of migraine or are sequelae triggered by recurrent migraine episodes.

## 5. Conclusion

In conclusion, this study elucidated RLS-associated brain functional changes, including ALFF, FC and topological properties in individuals with migraine and RLS. From the perspective of functional segregation and integration, the migraine with RLS exhibited significantly different local spontaneous neural activity, integration across distinct brain regions and functional networks features compared to migraine without RLS. Furthermore, migraine with RLS showed a notable correlations between RLS grading and ALFF in certain brain regions. Additionally, RLS severity is positively correlated with the angular gyrus betweenness centrality. This study enhances our understanding of RLS-related migraine and introduces a novel method for exploring the potential brain functional alterations in migraine with RLS.

## Data Availability

All data produced in the present study are available upon reasonable request to the authors

## Abbreviations

AAC: Anterior cingulate cortex
AAL: Anatomical automatic labeling
ALFF: Amplitude of low-frequency fluctuation
AUC: Area under the curve
BMI: Body mass index
BOLD: Blood oxygen level dependent
CSD: Cortical spreading depression
CTCD: Contrast-enhanced transcranial doppler
C_p_: Clustering coefficient
DICOM: Digital imaging and communications in medicine
DPABI: Data processing and analysis of brain imaging
DWI: Diffusion-weighted imaging
E_glob_: Global efficiency
E_loc_: Local efficiency
FC: Functional connectivity
FLAIR: Fluid attenuated inversion recovery
FOV: Field of view
FWMH: Full width half maximum
ICHD-3: International classification of headache disorders, 3rd edition
L_p_: Characteristic shortest path length
MCA: Middle cerebral artery
MNI: Montreal neurological institute
MPFC: Medial prefrontal cortex
NIfTI: Neuroimaging informatics technology initiative
PAG: Periaqueductal grey
PFC: Prefrontal cortex
PFO: Patent foramen ovale
RACC: Rodent anterior cingulate cortex
RLS: Right-to-left shunt
ROIs: Regions of interest
Rs-fMRI: Resting-state functional magnetic resonance imaging
SPSS: Statistical product and service solutions
T1WI: T1-weighted images
T2W2: T2-weighted images
TE: Echo time
TR: Repetition time
VAS: Visual analog scale
WMH: White matter hyperintensity
γ: Normalized clustering coefficient
σ: Small-worldness
λ: Normalized characteristic shortest path length

## Author contributions

WC, NW and JY conceived this study; WC contributed to the draft, and analyzed the data. WC, LJ, JZ, and HZ contributed to the collection of clinical and fMRI data. JY and NW strictly revised the manuscript. All authors have read and agreed to the published version of the manuscript.

## Acknowledgements

We would like to thank all the medical staff of the Department of Neurology at Shanghai Sixth People’s Hospital Affiliated to Shanghai Jiao Tong University School of Medicine.

## Availability of data and materials

The datasets analyzed during the current study are available from the corresponding author on reasonable request.

## Ethical approval

The study was carried out, according to the 1964 Declaration of Helsinki and its later amendments and according to local ethical guidelines.

## Consent for publication

All authors consent for the publication

## Reference

Arnold M (2018) Headache classification committee of the international headache society (IHS) the international classification of headache disordersJ. Cephalalgia 38(1): 1–211

Buse DC, Fanning KM, Reed ML et al (2019) Life with migraine: effects on relationships, career, and finances from the chronic migraine epidemiology and outcomes (CaMEO) study. Headache: The Journal of Head and Face Pain 59(8): 1286–1299

Chądzyński P, Kacprzak A, Domitrz W et al (2019) Migraine headache facilitators in a population of Polish women and their association with migraine occurrence - preliminary results. Neurol Neurochir Pol 53(5):377–383

Moon HJ, Seo JG, Park SP (2017) Perceived stress in patients with migraine: a case-control study. J Headache Pain 18(1):73

Fernández-de-Las-Peñas C, Fernández-Muñoz JJ, Palacios-Ceña M et al (2017) Sleep disturbances in tension-type headache and migraine. Ther Adv Neurol Disord 1:1756285617745444

Domitrz I, Cegielska J (2022) Magnesium as an Important Factor in the Pathogenesis and Treatment of Migraine-From Theory to Practice. Nutrients 14(5):1089

Goadsby PJ, Holland PR (2019) An Update: Pathophysiology of Migraine. Neurol Clin 37(4):651–671

Holland PR, Barloese M, Fahrenkrug J (2018) PACAP in hypothalamic regulation of sleep and circadian rhythm: importance for headache. J Headache Pain 19(1):20

Kim KM, Lee DH, Lee EJ et al (2019) Self-reported insomnia as a marker for anxiety and depression among migraineurs: a population-based cross-sectional study. Sci Rep 9(1):19608

Stanyer EC, Creeney H, Nesbitt AD et al. (2021) Subjective Sleep Quality and Sleep Architecture in Patients With Migraine: A Meta-analysis. Neurology 97(16):e1620–e1631

Wen Z, He M, Peng C et al (2019) Metabolomics and 16S rRNA Gene Sequencing Analyses of Changes in the Intestinal Flora and Biomarkers Induced by Gastrodia-Uncaria Treatment in a Rat Model of Chronic Migraine. Front Pharmacol 10:1425

Arzani M, Jahromi SR, Ghorbani Z et al (2020) Gut-brain Axis and migraine headache: a comprehensive review. J Headache Pain 21(1):15

Warnock JK, Cohen LJ, Blumenthal H et al. (2017) Hormone-Related Migraine Headaches and Mood Disorders: Treatment with Estrogen Stabilization. Pharmacotherapy 37(1):120–128

Hautakangas H, Winsvold BS, Ruotsalainen SE et al (2022) Genome-wide analysis of 102,084 migraine cases identifies 123 risk loci and subtype-specific risk alleles. Nat Genet 54(2):152–160

Sutherland HG, Albury CL, Griffiths LR (2019) Advances in genetics of migraine. J Headache Pain 20(1):72.

Zhao QX, Liu R, Zhou J et al (2021) Prevalence and grade of RLS in migraine: A prospective study of 251 migraineurs by synchronous test of c-TTE and c-TCD. Medicine (Baltimore) 100(4):e24175

Takagi H, Umemoto T, ALICE (All-Literature Investigation of Cardiovascular Evidence) Group (2016) A meta-analysis of case-control studies of the association of migraine and patent foramen ovale. J Cardiol 67(6):493–503

Liu K, Wang BZ, Hao Y et al (2020) The Correlation Between Migraine and Patent Foramen Ovale. Front Neurol 543485:1–12

Zhang YS, Jiang SL,, Zhu, et al (2021) Chinese expert guidelines for the prevention of patent foramen ovale-associated stroke. Chinese Heart Journal 33:1–10

Del Sette M, Angeli S, Leandri M et al (1998) Migraine with aura and right-to-left shunt on transcranial Doppler: a case-control study. Cerebrovasc Dis 8(6):327–330

Lip PZ, Lip GY (2014) Patent foramen ovale and migraine attacks: a systematic review. Am J Med 127: 411–420

Tian DC, Wang H, Chen W et al (2019) Meta-analysis of white matter lesions and patent foramen ovale in migraine. Neural Injury and Functional Reconstruction 14: 236–240+252

Zhao Q, Liu R, Zhou J et al (2021) Prevalence and grade of RLS in migraine: A prospective study of 251 migraineurs by synchronous test of c-TTE and c-TCD. Medicine (Baltimore) 100: e24175:1–e24175:7

Tang Y, Peng A, Peng B et al (2022) Association between patent foramen ovale and migraine without aura: a community-based cross-sectional study in China. BMJ Open 12: e056937:1-e056937:7

Dalla Volta G, Guindani M, Zavarise P et al (2005) Prevalence of patent foramen ovale in a large series of patients with migraine with aura, migraine without aura and cluster headache, and relationship with clinical phenotype. J Headache Pain 6(4):328–330

Wilmshurst P, Nightingale S (2006) The role of cardiac and pulmonary pathology in migraine: a hypothesis. Headache 46: 429–434

Ashina M, Hansen JM, Do TP et al (2019) Migraine and the trigeminovascular system-40 years and counting. Lancet Neurol 18: 795–804

Sevgi EB, Erdener SE, Demirci M et al (2012) Paradoxical air microembolism induces cerebral bioelectrical abnormalities and occasionally headache in patent foramen ovale patients with migraine. J Am Heart Assoc 1(6):e001735

Cao W, Shen Y, Zhong J et al (2022) The Patent Foramen Ovale and Migraine: Associated Mechanisms and Perspectives from MRI Evidence. Brain Sci 12(7):941

Park HK, Lee SY, Kim SE et al (2011) Small deep white matter lesions are associated with right-to-left shunts in migraineurs. J Neurol 258: 427–433

Yoon GJ, Kim JT, Chang J et al (2012). Right-to-left shunts as a cause of juxtacortical spots in patients with migraine. Eur J Neurol 19: 1086–1092

Iwasaki A, Suzuki K, Takekawa H et al (2017) The relationship between right-to-left shunt and brain white matter lesions in Japanese patients with migraine: a single center study. J Headache Pain 18(3):1–6

Vakamudi K, Trapp C, Talaat K et al (2012) Real-Time Resting-State Functional Magnetic Resonance Imaging Using Averaged Sliding Windows with Partial Correlations and Regression of Confounding Signals. Brain Connect 10(8):448–463

Zang YF, He Y, Zhu CZ et al (2012) Altered baseline brain activity in children with ADHD revealed by resting-state functional MRI Brain Dev 29(2):83–91

Wang N, Zeng W, Chen Let (2013) SACICA: a sparse approximation coefficient-based ICA model for functional magnetic resonance imaging data analysis. Journal of neuroscience methods 216(1):49–61

Wang N, Zeng W, Chen L (2012) A Fast-FENICA method on resting state fMRI data. Journal of Neuroscience Methods 209(1): 1–12

Tang XY, Zeng WM, Wang N Z et al (2017) A novel layered data reduction mechanism for clustering fMRI data. Biomedical Signal Processing and Control 33: 48–65

Wang Z, Li Y, Childress AR et al (2014) Brain entropy mapping using fMRI. PloS one 9(3): e89948

Lv H, Wang Z, Tong E et al (2018) Resting-State Functional MRI: Everything That Nonexperts Have Always Wanted to Know. AJNR Am J Neuroradiol 39(8):1390–1399

Zou QH, Zhu CZ, Yang Y et al (2008) An improved approach to detection of amplitude of low-frequency fluctuation (ALFF) for resting-state fMRI: fractional ALFF. Journal of neuroscience methods 172(1): 137–141

Yu Q, Cai Z, Li C et al (2021) A novel Spectrum contrast mapping method for functional magnetic resonance imaging data analysis. Frontiers in Human Neuroscience 15: 739668

Wang N, Chang C, Zeng W et al (2017) A novel feature-map based ICA model for identifying the individual, intra/inter-group brain networks across multiple fMRI datasets. Frontiers in neuroscience 11:510

Wang N, Zeng W, Shi Y et al (2015). WASICA: An effective wavelet-shrinkage based ICA model for brain fMRI data analysis. Journal of neuroscience methods 246: 75–96

Tang X, Zeng W, Wang N et al (2015). An adaptive RV measure based fuzzy weighting subspace clustering (ARV-FWSC) for fMRI data analysis. Biomedical Signal Processing and Control 22: 146–154

Tang XY, Zeng WM, Wang NZ et al (2017). A novel layered data reduction mechanism for clustering fMRI data. Biomedical Signal Processing and Control 33: 48–65

Wang N, Wu H, Xu M et al (2018). Occupational functional plasticity revealed by brain entropy: A resting-state fMRI study of seafarers. Human brain mapping 39(7): 2997–3004

Yan H, Wu H, Chen Y et al (2022). Dynamical complexity fingerprints of occupation-dependent brain functional networks in professional seafarers. Frontiers in Neuroscience 16: 830808

Sporns O (2018). Graph theory methods: applications in brain networks. Dialogues in clinical neuroscience 20(2): 111–121

Bullmore E, Sporns O (2009). Complex brain networks: graph theoretical analysis of structural and functional systems. Nature reviews neuroscience 10(3): 186–198

Messina R, Gollion C, Christensen RH et al (2022). Functional MRI in migraine. Curr Opin Neurol 35(3):328–335

Nie W, Zeng W, Yang J, et al (2021). Extraction and Analysis of Dynamic Functional Connectome Patterns in Migraine Sufferers: A Resting-State fMRI Study. Comput Math Methods Med. 2021:6614520

Lim M, Jassar H, Kim DJ et al (2021). Differential alteration of fMRI signal variability in the ascending trigeminal somatosensory and pain modulatory pathways in migraine J Headache Pain 22(1):4

Mungoven TJ, Marciszewski KK, Macefield VG et al (2022) Alterations in pain processing circuitries in episodic migraine. J Headache Pain 23(1):9

Zhang YS, Jiang SL, Zhu XY (2021) Chinese expert guidelines for the prevention of patent foramen ovale-associated stroke. Chin. Heart J 33:1–10

Yan C, Wang X, Zuo X et al (2016) DPABI: data processing & analysis for (resting-state) brain imaging. Neuroinformatics14(3):339–351

Yan C, Zang YF (2010) DPARSF: A MATLAB Toolbox for “Pipeline” Data Analysis of Resting-State fMRI. Front Syst Neurosci 4:13

Zuo XN, Di Martino A, Kelly C et al (2010) The oscillating brain: complex and reliable. Neuroimage 49(2):1432–1445

Biswal B, Yetkin FZ, Haughton VM et al (1995) Functional connectivity in the motor cortex of resting human brain using echo-planar MRI. Magn Reson Med 34(4):537–541

Tzourio-Mazoyer N, Landeau B, Papathanassiou D et al (2002) Automated anatomical labeling of activations in SPM using a macroscopic anatomical parcellation of the MNI MRI single-subject brain. Neuroimage 15(1):273–289

Rubinov M, Sporns O (2010) Complex network measures of brain connectivity: uses and interpretations. Neuroimage 52(3):1059–1069

Xia M, Wang J, He Y (2013) BrainNet Viewer: a network visualization tool for human brain connectomics. PloS one 8(7): e68910

Yan CG, Craddock RC, He Y et al (2013) Addressing head motion dependencies for small-world topologies in functional connectomics. Front Hum Neurosci 7:910

Yang H, Chen X, Chen ZB et al (2021) Disrupted intrinsic functional brain topology in patients with major depressive disorder. Mol Psychiatry 26(12):7363–7371

Zang YF, He Y, Zhu CZ et al (2007) Altered baseline brain activity in children with ADHD revealed by resting-state functional MRI. Brain Dev 29(2):83–91

Liu L, Hu X, Zhang Y et al (2020) Effect of Vestibular Rehabilitation on Spontaneous Brain Activity in Patients With Vestibular Migraine: A Resting-State Functional Magnetic Resonance Imaging Study. Front Hum Neurosci 14:227

Wang S, Wang H, Liu X et al (2023) A resting-state functional MRI study in patients with vestibular migraine during interictal period. Acta Neurol Belg 123(1):99–105

Zhe X, Tang M, Ai K et al (2023) Decreased ALFF and Functional Connectivity of the Thalamus in Vestibular Migraine Patients. Brain Sci 13(2):183

Khessali H, Mojadidi MK, Gevorgyan R et al (2012) The effect of patent foramen ovale closure on visual aura without headache or typical aura with migraine headache. JACC Cardiovasc Interv 5(6):682–687

Tu Y, Fu Z, Zeng F, et al (2019) Abnormal thalamocortical network dynamics in migraine. Neurology 92(23):e2706–e2716

Liu J, Zhao L, Li G et al (2012) Hierarchical alteration of brain structural and functional networks in female migraine sufferers. PLoS One 7(12):e51250

Baron R, Binder A, Wasner G (2010) Neuropathic pain: diagnosis, pathophysiological mechanisms, and treatment. Lancet Neurol 9(8):807–819

Lee GI, Neumeister MW (2020) Pain: Pathways and Physiology. Clin Plast Surg 47(2):173–180

Khan J, Asoom LIA, Sunni AA Genetics et al (2021) pathophysiology, diagnosis, treatment, management, and prevention of migraine. Biomed Pharmacother 139:111557

Ashina M, Hansen J M, Do T P et al (2019) Migraine and the trigeminovascular system—40 years and counting. The Lancet Neurology 18(8): 795–804

Li Z, Liu M, Lan L et al (2016) Altered periaqueductal gray resting state functional connectivity in migraine and the modulation effect of treatment. Sci Rep 6:20298

Li K, Liu L, Yin Q et al (2017) Abnormal rich club organization and impaired correlation between structural and functional connectivity in migraine sufferers. Brain Imaging Behav 11(2):526–540

Dai L, Zheng Q, Chen X et al (2021) Altered brain structural topological properties and its correlations with clinical characteristics in episodic migraine without aura. Neuroradiology 63(12):2099–2109

Labrakakis C (2023) The Role of the Insular Cortex in Pain. Int J Mol Sci 24(6):5736

Borsook D, Veggeberg R, Erpelding N et al (2016) The Insula: A “Hub of Activity” in Migraine. Neuroscientist 22(6):632–652

Li Q, Chen C, Gong T (2018) High-field MRS study of GABA+ in patients with migraine: response to levetiracetam treatment. Neuroreport 29(12):1007–1010

Kijima Y, Miller N, Noureddin N et al (2015) TCT-738 The Degree of Right-to-Left Shunt is Associated with Visual Aura Due to Migraine. J Am College Cardiol 66:B301

Dong L, Fan X, Fan Y et al (2023). Impairments to the multisensory integration brain regions during migraine chronification: correlation with the vestibular dysfunction. Front Mol Neurosci 16:1153641

de Pasquale F, Della Penna S, Snyder AZ et al (2012) A cortical core for dynamic integration of functional networks in the resting human brain. Neuron 74(4):753–764

